# Dose-response associations of device measured sleep regularity and duration with incident dementia in 82391 UK adults

**DOI:** 10.1101/2023.11.23.23298926

**Authors:** Wenxin Bian, Raaj K. Biswas, Matthew N. Ahmadi, Yu Sun Bin, Svetlana Postnova, Andrew J.K. Phillips, Nicolas A. Koemel, Jean-Philippe Chaput, Shantha M.W. Rajaratnam, Peter A Cistulli, Emmanuel Stamatakis

## Abstract

**Objectives:** To evaluate the associations of device-measured sleep duration and regularity with incident dementia, and to explore whether regular sleep might mitigate any association of sleep duration with dementia.

**Methods:** This population-based prospective cohort study of 82391 adults from the UK Biobank accelerometry subsample included adults aged 43 to 79 years old in England, Scotland, and Wales. Sleep duration (h/day) and Sleep Regularity Index (SRI, range 0-100) were calculated from the wrist-worn accelerometry data collected by participants over the course of one week. Cox proportional hazard models were used to estimate the hazard ratios (HRs) and assess the independent associations between sleep and incident dementia after adjustment for common demographic and contextual covariates.

**Results:** Over a mean follow-up of 7.9 years, during which 694 incident dementia cases occurred, there was a U-shaped association between sleep duration and incident dementia. Short sleep (<7 h) was associated with increased dementia risk, while long sleep (≥ 8h) was not significantly associated with dementia risk. The median sleep duration for short sleepers (<7 h) of 6.5 hours was associated with an HR of 1.19 (95% CI 1.01, 1.40) for incident dementia. Sleep regularity was negatively associated with dementia risk in a near-linear fashion. The sample median SRI of approximately 73, compared to the reference point of 51, was associated with an HR of 0.76 (95%CI 0.61, 0.94). The SRI value where the risk reduction was 50% of the maximum observed of 66, was associated with an HR of 0.77 (95%CI 0.63, 0.95). Among individuals with sleep duration outside the optimal range (too short or too long), less regular sleep was associated with increased risk of dementia. Among those with optimal sleep duration (7-8h/day), there was no significant association between sleep regularity and dementia risk. Compared to the reference point (SRI: 51), an SRI value of 62 for non-optimal sleepers was associated with a 25% reduction in risk for dementia (HR: 0.75; 95% CI 0.63, 0.90).

**Conclusions:** A regular sleep pattern may mitigate some adverse effects of inadequate sleep duration, suggesting that interventions aimed at improving sleep regularity may be a suitable option for people not able to achieve the recommended hours of sleep.

## INTRODUCTION

Dementia is a significant public health issue worldwide and a major non-communicable disease (NCD) [1]. The aging population has led to a growing concern for cognitive decline and dementia, with the World Health Organization (WHO) projecting a 141 million of affected people by 2050 [2, 3]. While dementia shares common behavioural risk factors with other NCDs, such as physical activity (PA), diet, sleep, and mental health, relatively little is known about the relationship between sleep and dementia [4]. Sleep plays a fundamental role in human physiology and is closely linked to numerous aspects of cognitive and physical well-being [5]. Despite its well-established importance for health, our understanding of how specific relationships between sleep and dementia is still limited. [6]

Existing studies have identified connections between sleep duration and health outcomes. A systematic review examining the relationship between sleep duration and a wide range of health outcomes in adults revealed that 7 to 8 hours of sleep per day exhibited the most favourable associations with health [7]. However, 96% of the studies used subjective sleep duration assessments [7]. Other studies have shown that shorter and longer sleep duration were associated with an increased risk in dementia amongst middle-aged to older adults [8–13]. The majority of studies to date were dependent on questionnaire-based measurement of sleep characteristics, which are more imprecise than objective measures and have inherent biases such as social desirability and poor recall [14, 15].

There is a growing interest in how other dimensions of sleep, including sleep regularity, are associated with adverse health outcomes. A systematic review on a broad range of 14 health outcomes revealed that more irregular sleep is generally associated with negative health outcomes [16]. The majority of the reviewed studies focused on cardiometabolic health and very few focused on brain health. Based on a recent report by the US National Sleep Foundation (NSF) that reviewed 63 full text publications identified that increased sleep irregularity was associated with adverse health and performance outcomes (e.g., mortality, cognitive performance, metabolic indicators, mental health, etc.) [17]. However, it remains unclear whether the adverse health effects of irregular sleep are independent of other sleep characteristics, such as sleep duration. Studies have shown that sleep duration can affect the association between sleep factors and their impact on health outcomes. Hence, understanding whether healthy sleep duration can mitigate the adverse health effects of an irregular sleep, and vice versa, is critical for public health and may offer practical means to improve health outcomes for individuals with unhealthy sleep patterns. Furthermore, those with inadequate sleep duration over weekdays may extend their sleep over weekends (“catch-up sleep”) to recover sleep debt, but this results in irregular sleep. Therefore, studies examining the association between sleep duration and regularity are critically important.

To date, no study has investigated whether increasing sleep regularity mitigates the adverse health effects of inadequate sleep duration on dementia. Thus, the aim of this study was to investigate the associations between device-measured sleep duration and regularity with incident dementia in adults. A secondary aim was to understand whether healthy sleep regularity might mitigate any deleterious association of sleep duration with dementia. We hypothesized that short and long sleep durations would be associated with incident dementia, and regularity in sleep timing would help mitigate the deleterious association of sleep duration with dementia.

## METHODS

### Study participants and design

The UK Biobank enrolled over 500,000 participants between 2006 and 2010 and informed written consent was provided [18]. The ethical approval was completed by the UK National Health service (NHS) and National Research Ethics Service for the UK (No. 11/NW/0382). Participants in the current study were drawn from the UK Biobank accelerometry sub-study, a prospective cohort of 103,104 participants aged 43 to 79 years. These participants were asked to wear a wrist accelerometer (Axivity AX3) on their dominant wrist to record daily rest–activity patterns for one week during June 2013 to January 2016 [19]. Participants with sufficient data from wrist accelerometers and non-missing information on all covariates were included. Accelerometer data were deemed sufficient if the participant wore the wearable device for at least three valid days (a minimum of 16 hours per day), with at least one of those days being a weekend day. Non-wear periods and the distinction between non-wear and sleep periods were identified following standardized procedures [20]. We defined the accelerometry baseline period as the follow-up time onset [21]. We excluded participants with diagnosed dementia prior to accelerometer wear.

### Sleep assessment

Sleep duration was calculated using a validated algorithm using relative changes in wrist tilt angle between successive 5 second windows [20, 22, 23]. For each interval of 5 seconds, the average of the estimated wrist tilt angle was calculated and served as an input for the algorithm. Sleep periods were those with a tilt angle change of less than 5 degrees for 5 minutes or longer. We used the Sleep Regularity Index (SRI) to assess participants’ sleep regularity [19, 24]. The SRI is a novel sleep indicator that quantifies sleep consistency by capturing day-to-day variation in sleep/wake patterns. We calculated SRI using the approach described in the literature [19]. The SRI is scaled from 0 to 100, where a greater value indicates a higher consistency in sleep patterns. It is more advantageous compared to other measures of regularity in sleep timing by assessing sleep-wake patterns between successive days without assuming any specific structure of sleep, making it applicable to populations with highly fragmented sleep or naps [19, 25].

### Outcome ascertainment

Incident dementia cases were obtained through data linkage with routinely-collected, coded national hospital admission, primary care and mortality data followed up to 30 November 2022 [26]. Inpatient hospitalization data were provided by the Hospital Episode Statistics for England, Scottish Morbidity Record and Patient Episode Database for Wales. Primary care data were obtained through collaborations with data providers. Death information was obtained from the National Health Service (NHS) Digital of England and Wales, and NHS Central Register and National Records of Scotland. Across all outcome event sources, the earliest date of dementia recorded was considered the date of diagnosis. We identified incident cases of dementia using previously validated ICD-10 (International Classification of Diseases) primary and secondary diagnoses provided in **Supplementary Table 1** [27].

### Covariates

We selected covariates based on previous literature [21, 23, 26, 28]. These included common demographic and contextual covariates such as age (years), sex (male, female), ethnicity (white, non-white), Body Mass Index (BMI) (continuous), fruit and vegetable consumption (servings per day), smoking status (current, previous, never), alcohol consumption (units per day), highest attained education level (A/AS level, College, CSE, NVQ/HND/HNC, O level, other), coffee consumption (cups per day), self-reported sleep problems including sleeplessness and insomnia (never/rarely, sometimes, usually), employment status (employed in day shift, night shift, no shift, not in workforce), device-based physical activity level (min/day) and sedentary behavior (h/day).

### Statistical analyses

In our study, the range of sleep duration and SRI values were capped at the 97.5 percentile to minimize the influence of sparse data. Cox proportional hazard models were used to assess the independent association between sleep duration and regularity, and the risk of developing dementia. We excluded participants with existing dementia to avoid reverse causation.

We carried out continuous dose-response analyses to investigate the potential linear and non-linear relationships between the exposures and the risk of incident dementia. Stratified analysis was used to explore the potential modification effect of sleep duration on sleep regularity. In the main analysis, we used the NSF sleep duration recommendation report for older adults [29], to stratify sleep duration into two groups, optimal (≥ 7 and < 8 h) and non-optimal (< 7h and ≥ 8h) sleepers. For additional analysis, we stratified sleep duration into three groups, short (< 7h), adequate (≥ 7 and < 8 h), long (≥ 8h) [29].

Based on the exposure distribution in all dose-response analyses, we placed the knots at the 10th, 50th, and 90th percentiles. The Wald test was used to assess non-linearity. For sleep duration, which we expected to have a U-shaped relationship, we first set the reference point to the lowest sleep data point. We estimated the optimal dose, which refers to the specific exposure value at which we observed the maximum significant risk reduction. We then set the reference point to the optimal dose for better graphical presentation. For sleep regularity, which had a monotonic relationship with the outcome, we also estimated the “minimum dose” as the exposure value at which the risk reduction was 50% of the maximum risk reduction observed [26].

Proportional hazards assumptions were tested using the Schoenfeld residuals to verify if the effect of the exposures on the hazard rate is constant over time. Models were adjusted for all covariates. All analyses were conducted in R version 4.3.0 and models were fitted using rms (version 6.7.0) and survival (version 3.5.5) packages.

### Sensitivity and additional analyses

We conducted sensitivity analyses: 1) excluding participants who developed dementia during the first two years of follow-up, and 2) excluding participants who reported poor self-rated health to account for the potential influence of reverse causation [30]. We also performed a sensitivity analysis excluding all participants who reported day or night shift work to remove the potential work schedule bias. We examined the association of sleep regularity with incident dementia modified by three groups of sleep duration (short, adequate, long).

Acknowledging the potential for loss of information associated with categorizing continuous variables, we conducted a categorical dose-response analysis in the **Supplementary** Figures 11 and 12. We categorized sleep regularity into three distinct groups based on tertiles.

## RESULTS

### Sample and events

Our analytic sample consisted of 82,391 participants with a mean (SD) age of 62.4 (7.7) years old; 55.9% were female, and 93.6% were White. Among the participants, a total of 783 cases of incident dementia were recorded during a mean follow-up time of 7.9 (1.0) years. Following reductions for data sparsity and outliers by truncating sleep exposure duration and regularity, we observed 694 and 729 incident dementia events in the analyses for sleep duration and regularity, respectively. Detailed characteristics of participants stratified by sleep duration and sleep regularity are presented in **Table 1** and **Supplementary Table 2**, respectively. A significant portion of participants (39.2%) had adequate sleep, followed by those having long (37.7%) or short sleep (23.0%).

**Table 1:** Participants baseline characteristics by sleep duration (n=82391).

| Characteristics | Overall | Short | Adequate | Long |
| --- | --- | --- | --- | --- |
| n | 82391 | 18987 | 32330 | 31074 |
| Age, y (mean (SD)) | 62.4 (7.7) | 62.9 (7.8) | 62.0 (7.8) | 62.5 (7.7) |
| Sex = Male, n (%) | 36339 (44.1) | 9770 (51.5) | 14231 (44.0) | 12338 (39.7) |
| Ethnic = White, n (%) | 77088 (93.6) | 17221 (90.7) | 30304 (93.7) | 29563 (95.1) |
| BMI (mean (SD)) | 26.7 (4.5) | 27.7 (4.9) | 26.6 (4.4) | 26.3 (4.3) |
| Fruit and vegetable consumption<sup>1</sup> (mean (SD)) | 8.0 (4.5) | 8.0 (4.8) | 8.1 (4.5) | 8.0 (4.4) |
| Smoking, n (%) |  |  |  |  |
| Current | 5339 (6.5) | 1516 (8.0) | 2052 (6.3) | 1771 (5.7) |
| Never | 47134 (57.2) | 10216 (53.8) | 18727 (57.9) | 18191 (58.5) |
| Previous | 29918 (36.3) | 7255 (38.2) | 11551 (35.7) | 11112 (35.8) |
| Alcohol consumption<sup>2</sup> (mean (SD)) | 13.71 (15.4) | 13.70 (16.5) | 13.59 (14.8) | 13.84 (15.2) |
| Coffee intake, cups per day (mean (SD)) | 2.0 (2.0) | 2.1 (2.1) | 2.0 (2.0) | 1.9 (1.9) |
| Mental health issues<sup>3</sup> = Yes, n (%) | 27005 (32.8) | 6440 (33.9) | 10109 (31.3) | 10456 (33.6) |
| Education, n (%) |  |  |  |  |
| A/AS level | 10618 (12.9) | 2463 (13.0) | 4118 (12.7) | 4037 (13.0) |
| College | 35996 (43.7) | 8390 (44.2) | 14727 (45.6) | 12879 (41.4) |
| CSE | 3225 (3.9) | 722 (3.8) | 1199 (3.7) | 1304 (4.2) |
| NVQ/HND/HNC | 4583 (5.6) | 1130 (6.0) | 1702 (5.3) | 1751 (5.6) |
| O level | 16696 (20.3) | 3666 (19.3) | 6474 (20.0) | 6556 (21.1) |
| Other | 11273 (13.7) | 2616 (13.8) | 4110 (12.7) | 4547 (14.6) |
| Sleeplessness and Insomnia<sup>4</sup> (%) |  |  |  |  |
| Never/rarely | 20515 (24.9) | 4729 (24.9) | 8533 (26.4) | 7253 (23.3) |
| Sometimes | 38967 (47.3) | 8491 (44.7) | 15343 (47.5) | 15133 (48.7) |
| Usually | 22909 (27.8) | 5767 (30.4) | 8454 (26.1) | 8688 (28.0) |
| Employment shift, n (%) |  |  |  |  |
| Employed in day shift work | 3285 (4.0) | 855 (4.5) | 1283 (4.0) | 1147 (3.7) |
| Employed in night shift work | 2936 (3.6) | 974 (5.1) | 1041 (3.2) | 921 (3.0) |
| Employed not in shift work | 40817 (49.5) | 8911 (46.9) | 16935 (52.4) | 14971 (48.2) |
| Retired/not in workforce | 35353 (42.9) | 8247 (43.4) | 13071 (40.4) | 14035 (45.2) |
| Lpa, mins per day (mean (SD)) | 118.4 (62.5) | 123.4 (65.2) | 121.0 (63.4) | 112.7 (59.3) |
| Mpa, mins per day (mean (SD)) | 34.8 (28.2) | 34.6 (28.9) | 35.9 (28.6) | 33.7 (27.2) |
| Vpa, mins per day (mean (SD)) | 5.3 (6.3) | 5.1 (6.4) | 5.6 (6.5) | 5.1 (5.9) |
| Sedentary Behaviour, mins per day (mean (SD)) | 715.4 (106.1) | 782.7 (104.6) | 716.3 (96.6) | 673.4 (94.6) |
| Dementia = Yes (%) | 3952 (4.8) | 1178 (6.2) | 1393 (4.3) | 1381 (4.4) |

The column breakdown corresponds to sleep duration: short, 0~7h/day; adequate, 7~8h/day; long, 8+h/day. There were significant differences ( $p<0.05$ ) across sleep score groups in all the characteristics shown in the table. Values represent mean (SD) unless specified otherwise. A/AS level.; CSE, ; Higher National Diploma, ; IQR, interquartile range; National Vocational Qualification, ; O level, .
<sup>1</sup>Fruits and vegetable consumption is servings per day.
<sup>2</sup>Alcohol consumption: above guidelines are >14 units per week, where 1 unit = 8 g of ethanol.
<sup>3</sup>Had ever seen a doctor or psychiatrist for nerves, anxiety or depression.
<sup>4</sup>Had trouble falling asleep at night or wake up in the middle of the night.

### Independent association of sleep duration and regularity with incident dementia

Figures 1 and **2** show the continuous dose-response association of sleep duration and regularity with incident dementia. In multivariable-adjusted analyses, there was a U-shaped relationship between sleep duration and the risk of incident dementia (Figure 1). The dose-response curve shows a higher risk of dementia in both directions, with a steeper increase in risk as sleep duration decreases and the reference point set at the optimal dose, i.e., lowest HR for dementia. However, the association between long sleep duration and dementia risk did not reach statistical significance (Figure 1). The optimal dose at which we set our reference point to was at 7.96 hours sleep per day. Sleep regularity and incident dementia exhibited a monotonic relationship, with the risk reduction in incident dementia declining with increasing sleep regularity for the whole population (Figure 2). Based on the predicted risk of incident dementia in the Cox proportional hazards model, the maximum risk reduction of 46% (HR: 0.54 [0.40-0.73]) was observed at the highest measured SRI value of 94.2, compared to the reference point at the lowest SRI value of 51.5. The minimum SRI dose, at which the risk reduction was 50% of the maximum risk reduction observed for sleep regularity was 66.1, corresponding to an HR of 0.77 (95%CI 0.63, 0.95). The median SRI dose was 72.9, corresponding to an HR of 0.76 (95%CI 0.61,0.94). Both irregular sleep and inadequate sleep were associated with higher dementia risk.

**Figure 1:**
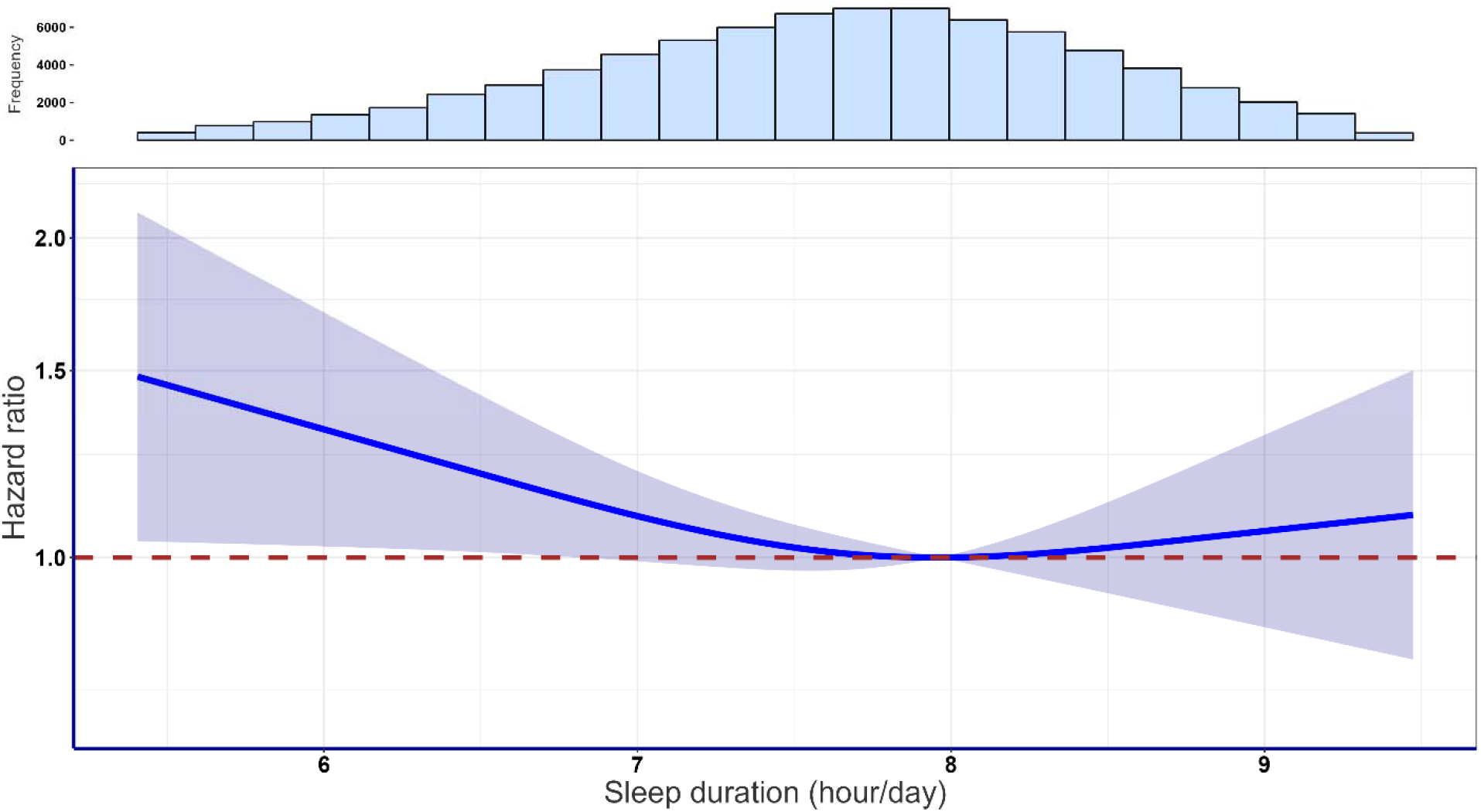
Association of sleep duration with incident dementia (n=78256, 694 events). Dose-response curves showing incident dementia HR associated with increasing daily sleep duration. Reference point set to the optimal data point (7.9 hrs of sleep/day). Adjusted for age, sex, ethnic, BMI, fruit and vegetable consumption, smoking, alcohol consumption, coffee consumption, mental health issue, sleeplessness/insomnia, education, shiftwork, LPA, MVPA and Sedentary behaviour. Data are shown for n=78256 with 694 events and with a mean follow-up of 7.9 (1.0) years.

**Figure 2:**
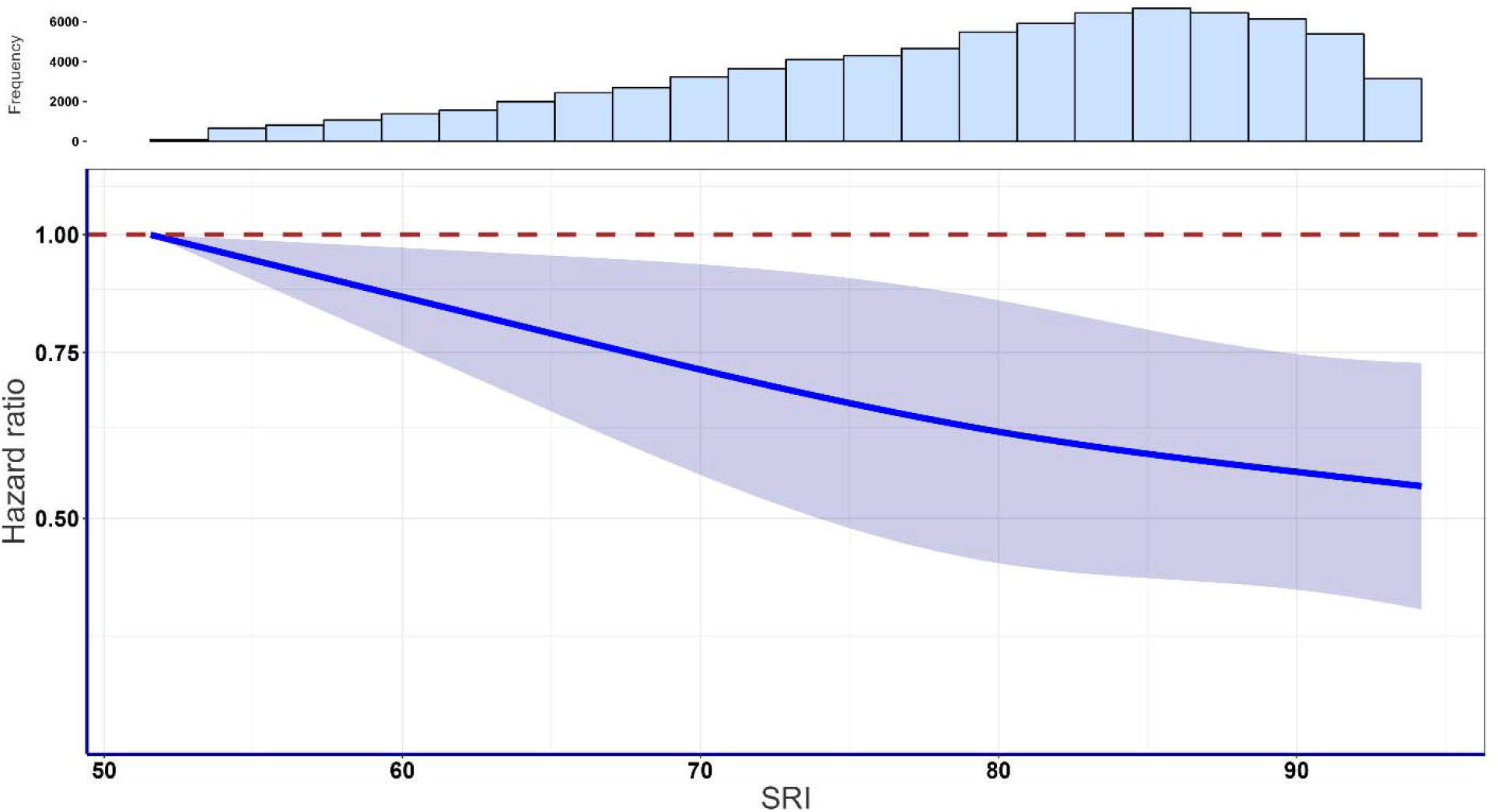
Association of sleep regularity with incident dementia (n=78262, 729 events). Dose-response curves showing incident dementia HR associated with increasing SRI. [10] Reference point set to the lowest data point (SRI=51.5). Adjusted for age, sex, ethnic, BMI, fruit and vegetable consumption, smoking, alcohol consumption, coffee consumption, mental health issue, sleeplessness/insomnia, education, shiftwork, LPA, MVPA, sedentary behaviour. Data are shown for n=78262 with 729 events and with a mean follow-up of 7.9 (1.0) years.

### Associations of sleep regularity with incident dementia by sleep duration status

Figure 3 shows the dose-response relationship between sleep regularity and incident dementia modified by groups of sleep duration (optimal ≥ 7 and < 8 h, non-optimal (< 7h and > 8h). For non-optimal sleepers, we observed a significant inverse association between sleep regularity and incident dementia. The curve demonstrated a consistent descend in correspondence with the increase in SRI, after which started to stabilize at a risk reduction of around 50%. The minimum sleep regularity dose for non-optimal sleepers was 61.6, corresponding to a HR of 0.75 (95% CI 0.63, 0.90). The median dose was 72.9, corresponding to a HR of 0.65 (96%CI 0.50, 0.85). For optimal sleepers, the curve initially exhibited a relatively gentle slope of no effect, with little variation as SRI increases. The curve started to steadily descend at an accelerating rate after crossing the null line (HR=1) at an SRI value of 80.9. The minimum sleep regularity dose for optimal sleepers was 87.3, corresponding to a HR of 0.80 (95% CI 0.45, 1.41). However, the wide confidence interval of the curve suggests that the association between sleep regularity and incident dementia for optimal sleepers were not statistically significant (Figure 3). The statistical interaction test suggested that there was no interaction between sleep duration and sleep regularity (**Supplementary Table 3**).

**Figure 3:**
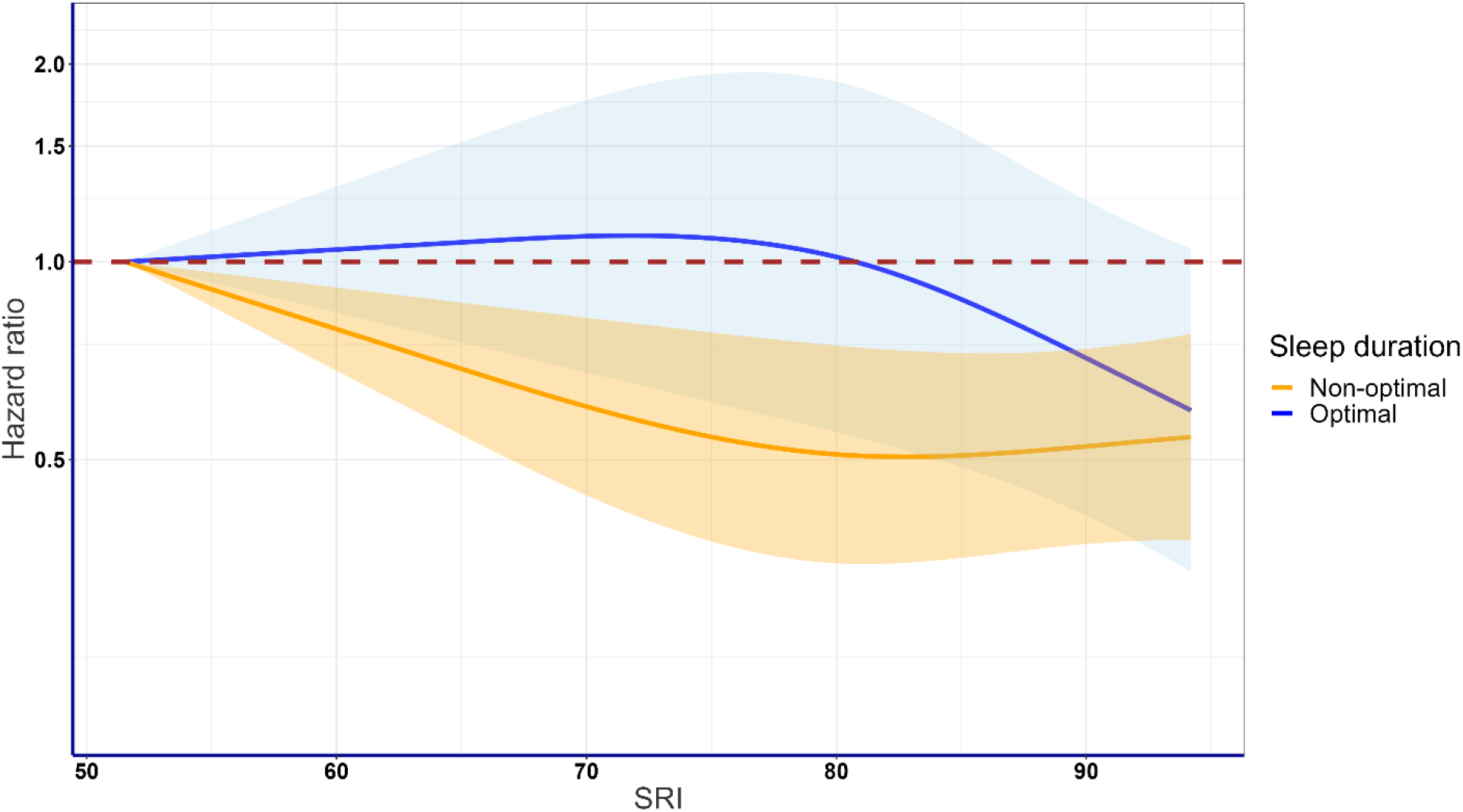
Association of sleep regularity with incident dementia stratified by sleep duration (n=78262, 729 events). Dose-response curves showing incident dementia HR associated with increasing SRI stratified by two groups of sleep duration (optimal ≥ 7 and < 8h/day, and non-optimal <7h/day and ≥ 8h/day). Reference point set to lowest data point (SRI=51.5). Adjusted for age, sex, ethnic, BMI, fruit and vegetable consumption, smoking, alcohol consumption, coffee consumption, mental health issue, sleeplessness/insomnia, education, shiftwork, LPA, MVPA and sedentary behaviour. Data are shown for n=78262 with 729 events and with a mean follow-up of 7.9 (1.0) years.

### Sensitivity & Additional analyses

Excluding participants who developed dementia within the first two years of follow-up (events=69) had minimal impact on the overall pattern and results, as shown in **Supplementary** Figures 5 to 7. The minimum SRI dose for sleep regularity in this group was 64.4 with a HR of 0.77 (95%CI 0.64, 0.93), while the minimum sleep regularity dose for non-optimal sleepers was 61.4, corresponding to a HR of 0.74 (95% CI 0.62, 0.88). Excluding participants with self-reported poor health (n=2056) also did not produce substantial changes in the results, as indicated in **Supplementary** Figure 8 to 10. The minimum SRI dose for sleep regularity in this group was 68.4 with a HR of 0.79 (95%CI 0.62, 1.01), while the minimum sleep regularity dose for non-optimal sleepers was 62.4, corresponding to a HR of 0.78 (95% CI 0.64, 0.95). Similarly, excluding participants employed in shift work (n=6221) had a minimal impact on the results (**Supplementary** Figure 2 to 4). The minimum SRI dose for sleep regularity was 66.1 with a HR of 0.77 (95%CI 0.63, 0.95). The minimum sleep regularity dose for non-optimal sleepers was 62.0, corresponding to a HR of 0.74 (95% CI 0.62, 0.89). Notably, the optimal dose at which the maximum significant risk reduction was observed remained consistent at 7.96 hours of sleep per day for all sensitivity analyses. No association was found between sleep regularity and the risk of incident dementia for individuals with an optimal sleeping duration. Continuous dose-response analysis with three groups of sleep duration (short, adequate, long) showed comparable inverse associations in the dose-response curves for the short and long categories in contrast to the adequate group (**Supplementary** Figure 1). Categorical analysis of sleep duration (**Supplementary** Figure 11) by three groups based on the literature shows an increased risk of incident dementia for participants with short sleep durations with HR 1.18 (0.97,1.43) compared to those with adequate sleep. There was no significant association between long sleep duration and risk of incident dementia with a HR of 0.95 (0.79,1.14). Categorical analysis of sleep regularity (**Supplementary** Figure 12) by population tertile-based groups produced results consistent with the main dose-response analysis results.

## DISCUSSION

To our knowledge, our study is the first to examine the association between sleep duration and sleep regularity and the risk of incident dementia. Our results suggest a U-shaped dose-response relationship of short sleep durations with incident dementia. More regular sleep patterns were associated with lower dementia risk in a dose response manner. The sample median SRI of approximately 73, representing the percentage probability of an individual being in the same sleep/wake state at any two time points (0 being random and 100 perfectly regular), was associated with an HR of 0.76 (95%CI 0.61, 0.94). Among participants with short and long sleep, the beneficial associations between sleep regularity and incident dementia were more pronounced than in those within the optimal sleep range.

Sleep duration demonstrated a U-shaped association with incident dementia. However, statistical significance was reached only in individuals with short sleep duration. Our findings are in line with previous literature that found short device-measured sleep duration to be associated with dementia, but not for long sleep duration [28]. Other recent meta-analyses of self-reported studies suggested a significant U-shaped association between sleep duration and dementia risk, with a sleep duration of around 7 hours being the most beneficial [31, 32]. Additionally, a meta-analysis of prospective cohort studies showed that there is a significant association between longer sleep duration and dementia risk [33]. However, one study suggested that the association between self-reported long sleep duration (>9h per night) and increased dementia risk might be due to reverse causation, as the association did not remain after exclusion of cases diagnosed within the first 5 or 10 years of follow-up [6].

Recent studies have indicated that sleep regularity is associated with a wide range of health outcomes, including metabolic health, obesity, mortality, mental health, etc. [34–39]. In the whole sample, our results indicated an inverse monotonic near-linear association between sleep regularity and incident dementia risk. Another wearables-based study examined the association between sleep-wake patterns and incident dementia [40]. It also revealed that greater variability in sleep duration was associated with increased risk of developing dementia, using the standard deviation of sleep duration as a measure of daily sleep consistency [40]. Our study used the SRI as a sleep regularity measurement, which captures variations in multiple dimensions of sleep patterns, covering not only sleep duration, but also sleep onset and offset, naps and sleep-wake state within the sleep window.

Our findings suggest that sleep regularity might help mitigate the deleterious association of sleep duration with dementia. It also suggests that sleep regularity may matters less when sleep duration is in the optimal range. Among participants with optimal sleep duration, sleep regularity showed less clear associations with incident dementia. For non-optimal sleepers defined as participants who sleep less than 7 hours or more than 8 hours per day, we observed a significant inverse association between sleep regularity and incident dementia. The decline in incident dementia risk was consistent in the first half of the SRI distribution, levelling off as the SRI value surpassed around 80. The median SRI value of approximately 72 was associated with a HR of 0.65 (95%CI 0.50, 0.85). A recent study using SRI scores from accelerometer data in the UK Biobank found that sleep regularity was more strongly associated with mortality risk than sleep duration [39]. For those with non-optimal sleep duration, maintaining regular sleep patterns could potentially mitigate the adverse impact of non-optimal sleep durations on the risk of developing dementia.

Experimental and epidemiological studies provide a plausible mechanism linking poor sleep patterns and Alzheimer’s disease (AD), which is the most common form of dementia [41, 42]. Sleep critically regulates AD-related proteins, such as amyloid-β (Aβ) and tau, preventing their accumulation resulting in aggregates critical in pathogenesis [43]. Epidemiological evidence using self-reported sleep duration demonstrated that short and long sleep duration were associated with greater Aβ burden [44]. Lack of sleep has specifically been linked to heightened production of Aβ at synapses during wakefulness compared to sleep, as well as a decrease in the clearance of Aβ from the brain’s interstitial space [45, 46]. We note that sleep regularity is an indirect approach to assessing the degree of alignment between sleep and circadian timing (and light-dark cycles), which is known to have biological relationships with dementia [47–50]. Experimental studies on mice have shown that circadian disruption negatively affects the Aβ dynamics and speeds up the build-up of amyloid plaques in the brain [41]. Given that studies have indicated the circadian system’s role in regulating amyloid-beta levels in the brain, it may partly explain the potential role of regular sleep in mitigating the adverse effects of unhealthy sleep duration.

The role of sleep regularity in dementia has not been investigated in a large cohort with objective data. Our study assessed concurrently two important sleep dimensions, duration and regularity, providing a comprehensive understanding of the multidimensional nature of sleep and its association with dementia. Other strengths include large sample size, objective sleep measurements, the use of a novel sleep regularity metric [19], and the performance of both continuous and categorical analyses. Our study had several limitations. The study acknowledges that not all potential confounders have been fully accounted for. The accelerometry data were collected 5.5 years after the baseline data collection, however covariates were relatively stable over time, with the exception of employment status [23, 51]. The response rate in the UK Biobank was 5.5% [52], however a previous study suggested that the association of lifestyle exposures and long term health outcomes is not materially influenced by poor sample representativeness [53]. Our results pertain to an older age group predominantly consisting of individuals from a homogeneous ethnic background. To enhance generalizability of our finding, further research with other diverse cross-cultural cohorts may be warranted.

## CONCLUSION

Sleep duration and regularity were associated with risk of incident dementia. Sleep regularity showed an inverse association with incident dementia among those with non-optimal sleep duration, while no significant association was observed among those with optimal sleep duration. In the presence of inadequate sleep duration, maintaining regular sleep timing may reduce the risk of incident dementia. Considering the challenges of behaviour change, future guidelines could place equal emphasis on sleep regularity and duration, to expand intervention options and personalised advice.

## Data Availability

The data that support the findings of this study are available from UK Biobank http://www.ukbiobank.ac.uk/. Restrictions apply to the availability of these data, which were used under application number 25813, and so are not publicly available. Data are available from the authors upon reasonable request and with permission of the UK Biobank.

## Acknowledgements

This research has been conducted using the UK Biobank resource under application number 25813. The authors would like to thank all the participants and professionals contributing to the UK Biobank. All information and materials in the manuscript are original and have not been submitted for publication elsewhere.

