## Supplementary Material for "Dose-response associations of device measured sleep regularity and duration with incident dementia in 82391 UK adults"

**Supplemental Table 1**: Dementia; ICD 10 classifications

| **Dementia type (ICD 10 classification)** | **Code** |
| --- | --- |
| Dementia in Alzheimer's disease | F00 |
| Dementia in Alzheimer's disease with early onset | F00.0 |
| Dementia in Alzheimer's disease with late onset | F00.1 |
| Dementia in Alzheimer's disease, atypical or mixed type | F00.2 |
| Dementia in Alzheimer's disease, unspecified | F00.9 |
| Alzheimer’s disease | G30 |
| Alzheimer’s disease with early onset | G30.0 |
| Alzheimer’s disease with late onset | G30.1 |
| Other Alzheimer's disease | G30.8 |
| Alzheimer's disease unspecified | G30.9 |
| Vascular dementia | F01 |
| Vascular dementia of acute onset | F01.0 |
| Multi-infarct dementia | F01.1 |
| Subcortical vascular dementia | F01.2 |
| Mixed cortical and sub-cortical vascular dementia | F01.3 |
| Other vascular dementia | F01.8 |
| Vascular dementia, unspecified | F01.9 |
| Binswanger's disease | I67.3 |
| Dementia in Picks disease | F02.0 |
| Circumscribed brain atrophy | G31.0 |
| Sporadic Creutzfeldt-Jakob disease | A81.0 |
| Dementia in Creutzfeldt-Jacob disease | F02.1 |
| Dementia in Huntington’s disease | F02.2 |
| Dementia in Parkinson’s disease | F02.3 |
| Dementia in HIV disease | F02.4 |
| Mental and behavioural disorders due to use of alcohol - amnesic syndrome | F10.6 |
| Dementia in other diseases classified elsewhere | F02 |
| Dementia in other specified diseases classified elsewhere | F02.8 |
| Unspecified dementia | F03 |
| Delirium superimposed on dementia | F05.1 |
| Senile degeneration of brain | G31.1 |
| Other specified degenerative diseases of nervous system | G31.8 |

| **Dementia type (Read version 2)** | **Code** |
| --- | --- |
| [X] Dementia in Alzheimer's disease | Eu00. |
| [X]Dementia in Alzheimer's disease with early onset | Eu000 |
| [X]Dementia in Alzheimer's disease with late onset | Eu001 |
| [X]Dementia in Alzheimer's disease, atypical or mixed type | Eu002 |
| [X]Dementia in Alzheimer's disease, unspecified | Eu00z |
| Alzheimer’s disease | F110. |
| Alzheimer’s disease with early onset | F1100 |
| Alzheimer’s disease with late onset | F1101 |
| Senile degeneration of brain | F112. |
| [X] Other Alzheimer's disease | Fyu30 |
| Multi-infarct dementia | E004. |
| Uncomplicated arteriosclerotic dementia | E0040 |
| Arteriosclerotic dementia with delirium | E0041 |
| Arteriosclerotic dementia with paranoia | E0042 |
| Arteriosclerotic dementia with depression | E0043 |
| Arteriosclerotic dementia NOS | E004z |
| [X]Vascular dementia | Eu01. |
| [X]Vascular dementia of acute onset | Eu010 |
| [X]Multi-infarct dementia | Eu011 |
| [X]Other vascular dementia | Eu01y |
| [X]Vascular dementia, unspecified | Eu01z |
| Cerebral degeneration due to cerebrovascular disease | F11x2 |
| Binswanger's disease | F21y2 |
| [X] Lewy body dementia | Eu025 |
| Lewy body disease | F116. |
| [X] Dementia in Picks disease | Eu020 |
| Pick's disease | F111. |
| Frontotemporal degeneration | F118. |
| Jakob-Creutzfeldt disease | A411. |
| Sporadic Creutzfeldt-Jakob disease | A4110 |
| Alcoholic dementia, NOS | E012. |
| Dementia in conditions EC | E041. |
| [X] Dementia in other diseases classified elsewhere | Eu02. |
| [X] Dementia in Creutzfeldt-Jacob disease | Eu021 |
| [X] Dementia in Huntington’s disease | Eu022 |
| [X] Dementia in Parkinson’s disease | Eu023 |
| [X] Dementia in HIV disease | Eu024 |
| [X]Dementia in other specified diseases classified elsewhere | Eu02y |
| [X]Mental and behavioural disorders due to use of alcohol:  amnesic syndrome | Eu106 |

| [X]Mental and behavioural disorders due to use of alcohol:  residual and late-onset psychotic disorder | Eu107 |
| --- | --- |
| Cerebral degeneration due to Jacob-Creutzfeldt disease | F11x7 |
| Cerebral degeneration due to Parkinson’s disease | F11x9 |
| Corticobasal degeneration | F11y2 |
| H/O: dementia | 1461. |
| Assessment of psychotic and behavioural symptoms of dementia | 38C13 |
| GDS level 4 - moderate cognitive decline | 3AE3. |
| GDS level 5 - moderately severe cognitive decline | 3AE4. |
| GDS level 6 - severe cognitive decline | 3AE5. |
| GDS level 7 - very severe cognitive decline | 3AE6. |
| Dementia monitoring | 66h.. |
| Dementia annual review | 6AB.. |
| Dementia medication review | 8BM02 |
| Shared care – prescribing drug for dementia | 8BM50 |
| Shared care – prescribing drug for dementia declined | 8BM60 |
| Antipsyc drug therapy dementia | 8BPa. |
| Dementia advance care plan | 8CMe0 |
| Review of dementia advance care plan | 8CMG2 |
| Dementia care plan | 8CMZ. |
| Dementia care plan agreed | 8CMZ0 |
| Dementia care plan reviewed | 8CMZ1 |
| Dementia care plan declined | 8CMZ2 |
| Dementia care plan review declined | 8CMZ3 |
| Dementia advance care plan agreed | 8CSA. |
| Referral to dementia care advisor | 8Hla. |
| Dementia adv care plan declnd | 8IAe0 |
| Dementia advance care plan review declined | 8IAe2 |
| Exception reporting: dementia quality indicators | 9hD.. |
| Excepted from dementia quality indicators: patient unsuitable | 9hD0. |
| Excepted from dementia quality indicators: informed dissent | 9hD1. |
| Dementia monitoring administration | 9Ou.. |
| Dementia monitoring first letter | 9Ou1. |
| Dementia monitoring second letter | 9Ou2. |
| Dementia monitoring third letter | 9Ou3. |
| Dementia monitoring verbal invite | 9Ou4. |
| Dementia monitoring telephone invite | 9Ou5. |
| Senile and presenile organic psychotic condition | E00.. |
| Uncomplicated senile dementia | E000. |
| Pre-senile dementia | E001. |
| Uncomplicated pre-senile dementia | E0010 |
| Pre-senile dementia with delirium | E0011 |
| Pre-senile dementia with paranoia | E0012 |
| Pre-senile dementia with depression | E0013 |
| Pre-senile dementia NOS | E001z |
| Senile dementia with depressive or paranoid features | E002. |
| Senile dementia with paranoia | E0020 |
| Senile dementia with depression | E0021 |
| Senile dementia with depressive or paranoid features NOS | E002z |
| Senile dementia with delirium | E003. |
| Drug induced dementia | E02y1 |
| [X]Sub-cortical vascular dementia | Eu012 |
| [X]Mixed cortical and sub-cortical vascular dementia | Eu013 |
| [X] Unspecified dementia | Eu02z |
| [X] Delirium superimposed on dementia | Eu041 |

**Supplementary Table 2**: Participants baseline characteristics by sleep regularity (n=82391).

|  | Overall | Irregular | Regular |
| --- | --- | --- | --- |
| n | 82391 | 17869 | 64522 |
| Age, y (mean (SD)) | 62.4 (7.7) | 62.7 (7.8) | 62.3 (7.7) |
| Sex = Male, n (%) | 36339 (44.1) | 7065 (39.5) | 29274 (45.4) |
| Ethnic = White, n (%) | 77088 (93.6) | 16572 (92.7) | 60516 (93.8) |
| BMI (mean (SD)) | 26.7 (4.5) | 27.3 (4.9) | 26.6 (4.4) |
| Fruit and vegetable consumption^1^ (mean (SD)) | 8.0 (4.5) | 8.0 (4.6) | 8.0 (4.5) |
| Smoking, n (%) |  |  |  |
| Current | 5339 (6.5) | 1490 (8.3) | 3849 (6.0) |
| Never | 47134 (57.2) | 9663 (54.1) | 37471 (58.1) |
| Previous | 29918 (36.3) | 6716 (37.6) | 23202 (36.0) |
| Alcohol consumption^2^ (mean (SD)) | 13.7 (15.4) | 13.8 (16.3) | 13.7 (15.1) |
| Coffee intake, cups per day (mean (SD)) | 2.0 (2.0) | 2.0 (2.0) | 2.0 (2.0) |
| Mental health issue^3^ = Yes, n (%) | 27005 (32.8) | 6589 (36.9) | 20416 (31.6) |
| Education, n (%) |  |  |  |
| A/AS level | 10618 (14.9) | 2278 (15.1) | 8340 (14.9) |
| College | 35996 (50.6) | 7257 (48.0) | 28739 (51.3) |
| CSE | 3225 (4.5) | 772 (5.1) | 2453 (4.4) |
| NVQ/HND/HNC | 4583 (6.4) | 1052 (7.0) | 3531 (6.3) |
| O level | 16696 (23.5) | 3766 (24.9) | 12930 (23.1) |
| Sleeplessness and insomnia^4^ (%) |  |  |  |
| Never/rarely | 20515 (24.9) | 3825 (21.4) | 16690 (25.9) |
| Sometimes | 38967 (47.3) | 8228 (46.0) | 30739 (47.6) |
| Usually | 22909 (27.8) | 5816 (32.5) | 17093 (26.5) |
| Employment shift, n (%) |  |  |  |
| Employed in day shift work | 3285 (4.0) | 799 (4.5) | 2486 (3.9) |
| Employed in night shift work | 2936 (3.6) | 999 (5.6) | 1937 (3.0) |
| Employed not in shift work | 40817 (49.5) | 8056 (45.1) | 32761 (50.8) |
| Retired/not in workforce | 35353 (42.9) | 8015 (44.9) | 27338 (42.4) |
| Lpa, mins per day (mean (SD)) | 118.4 (62.5) | 114.0 (60.6) | 119.7 (63.0) |
| Mpa, mins per day (mean (SD)) | 34.8 (28.2) | 31.2 (26.7) | 35.8 (28.5) |
| Vpa, mins per day (mean (SD)) | 5.3 (6.3) | 4.4 (5.6) | 5.5 (6.5) |
| Sedentary behaviour, mins per day (mean (SD)) | 715.4 (106.1) | 738.7 (111.2) | 709.0 (103.7) |
| Dementia = Yes (%) | 783 (1.0) | 239 (1.3) | 544 (0.8) |

The columns breakdown corresponds to sleep regularity: irregular, 0~70; regular, 70~100. There were significant differences (p<0.05) across sleep score groups in all the characteristics shown in the table. Values represent mean (SD) unless specified otherwise. A/AS level. ; CSE, ; Higher National Diploma, ; IQR, interquartile range; National Vocational Qualification, ; O level, .

^1^Fruits and vegetable consumption is servings per day.

^2^Alcohol consumption: above guidelines is >14 units per week, where 1 unit = 8 g of ethanol.

^3^Had ever seen a doctor or psychiatrist for nerves, anxiety or depression.

^4^Had trouble falling asleep at night or wake up in the middle of the night.

**Supplementary Figure 1**: Association of sleep regularity with incident dementia stratified by sleep duration (n=7826, 729 events).

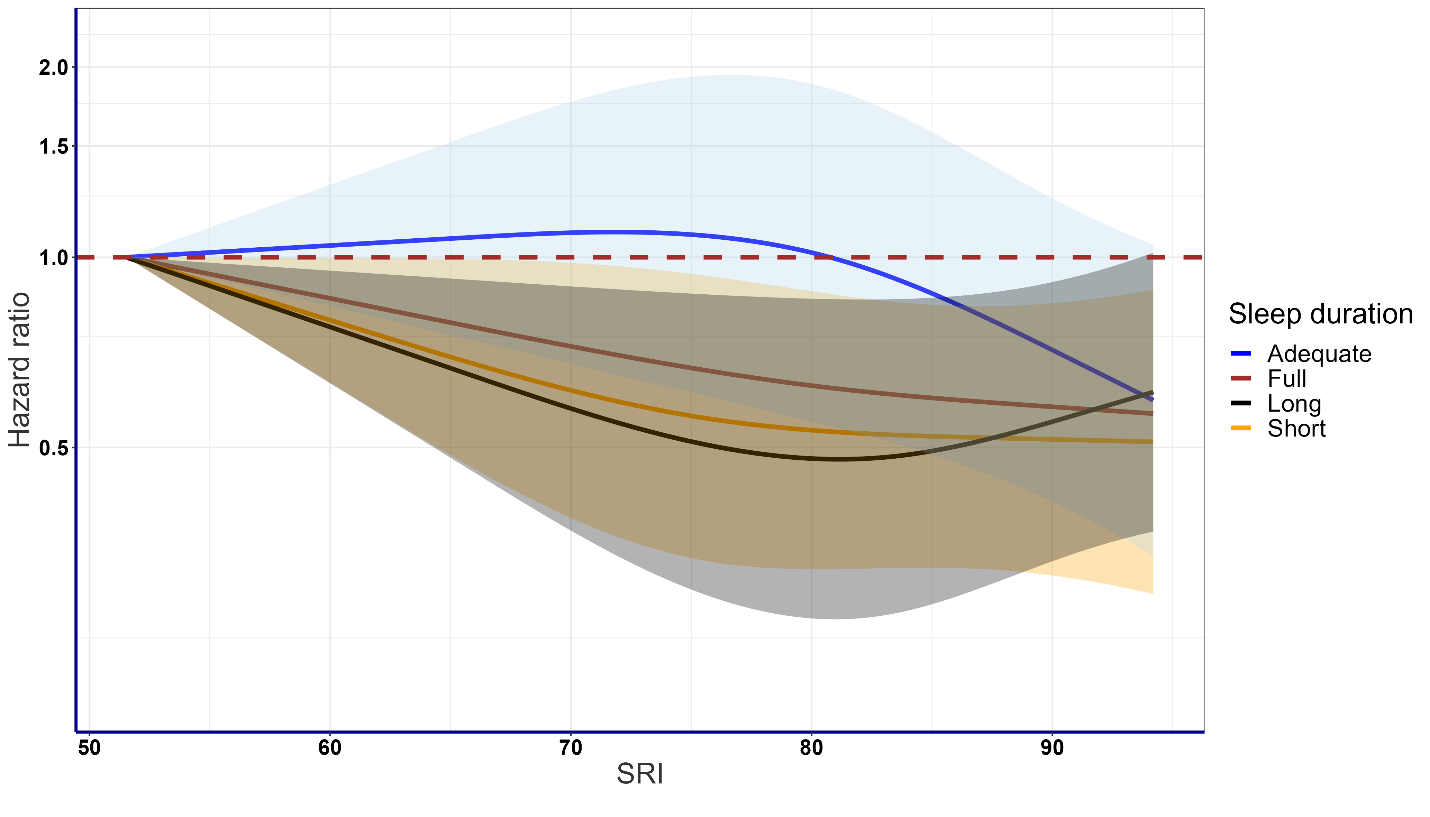

Dose-response curves showing incident dementia HR associated with increasing SRI stratified by three groups of sleep duration (adequate ≥ 7 and < 8h/day, short <7h/day and long ≥ 8h/day). A curve with the full dataset (i.e., without any stratification) is also provided. Reference point set to lowest data point (SRI=51.5). Adjusted for age, sex, ethnic, BMI, fruit and vegetable consumption, smoking, alcohol consumption, coffee consumption, mental health issue, sleeplessness/insomnia, education, shiftwork, LPA, MVPA, sedentary y behaviour. Data are shown for n=78262 with 729 events and with a mean follow-up of 7.9 (1.0) years.

**Supplementary Figure 2**: Association of sleep duration with incident dementia with participants employed in shift work excluded (n=72355, 666 events).

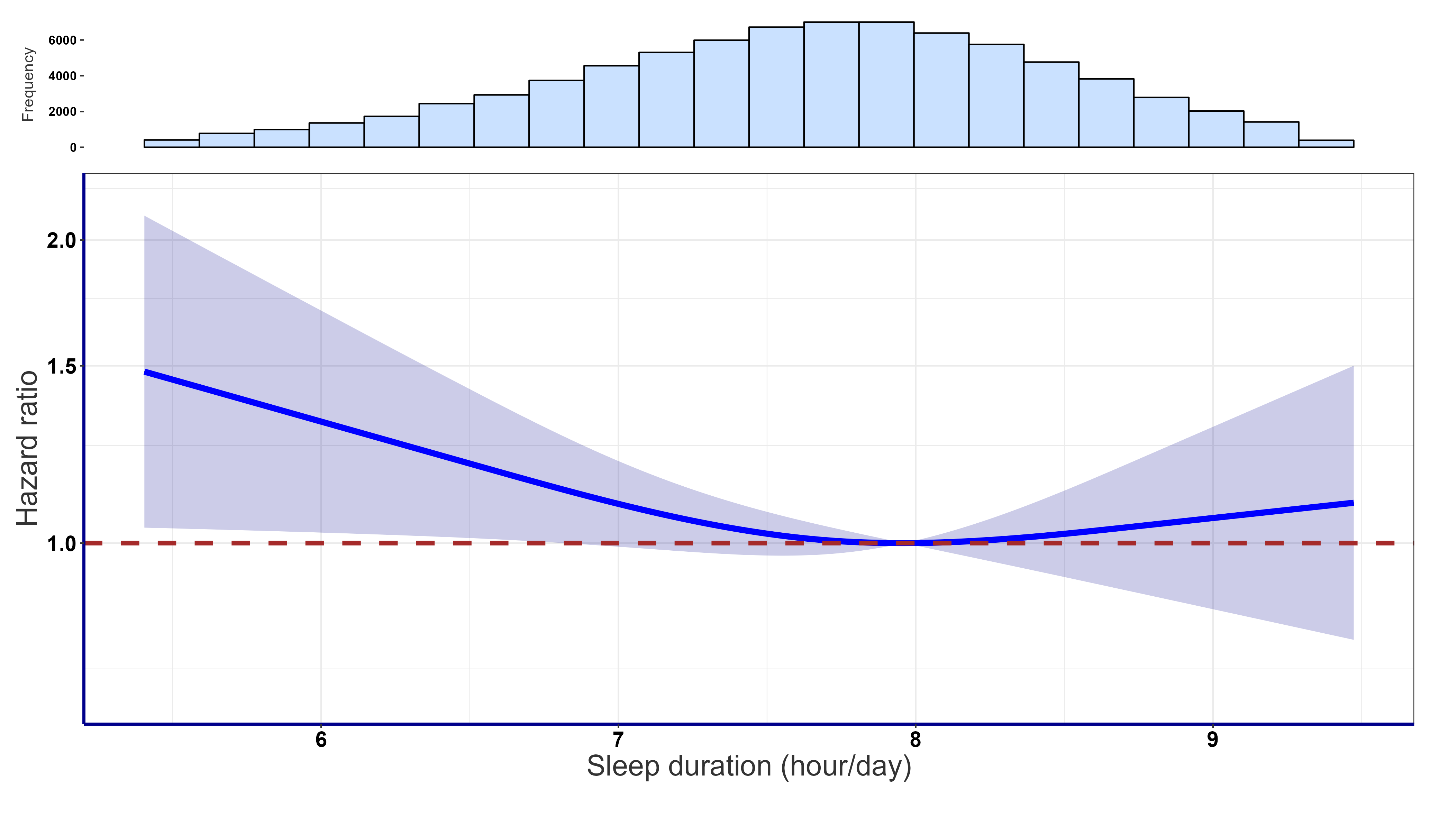

Dose-response curves showing incident dementia HR associated with increasing daily sleep duration. Reference point set to the optimal data point (7.9 hrs of sleep/day). Adjusted for age, sex, ethnic, BMI, fruit and vegetable consumption, smoking, alcohol consumption, coffee consumption, mental health issue, sleeplessness/insomnia, education, shiftwork, LPA, MVPA, Sedentary behaviour. Data are shown for n=72355 with 666 events and with a mean follow-up of 7.9 (1.0) years.

**Supplementary Figure 3**: Association of sleep regularity with incident dementia with participants employed in shift work excluded (n=72347, 703 events).

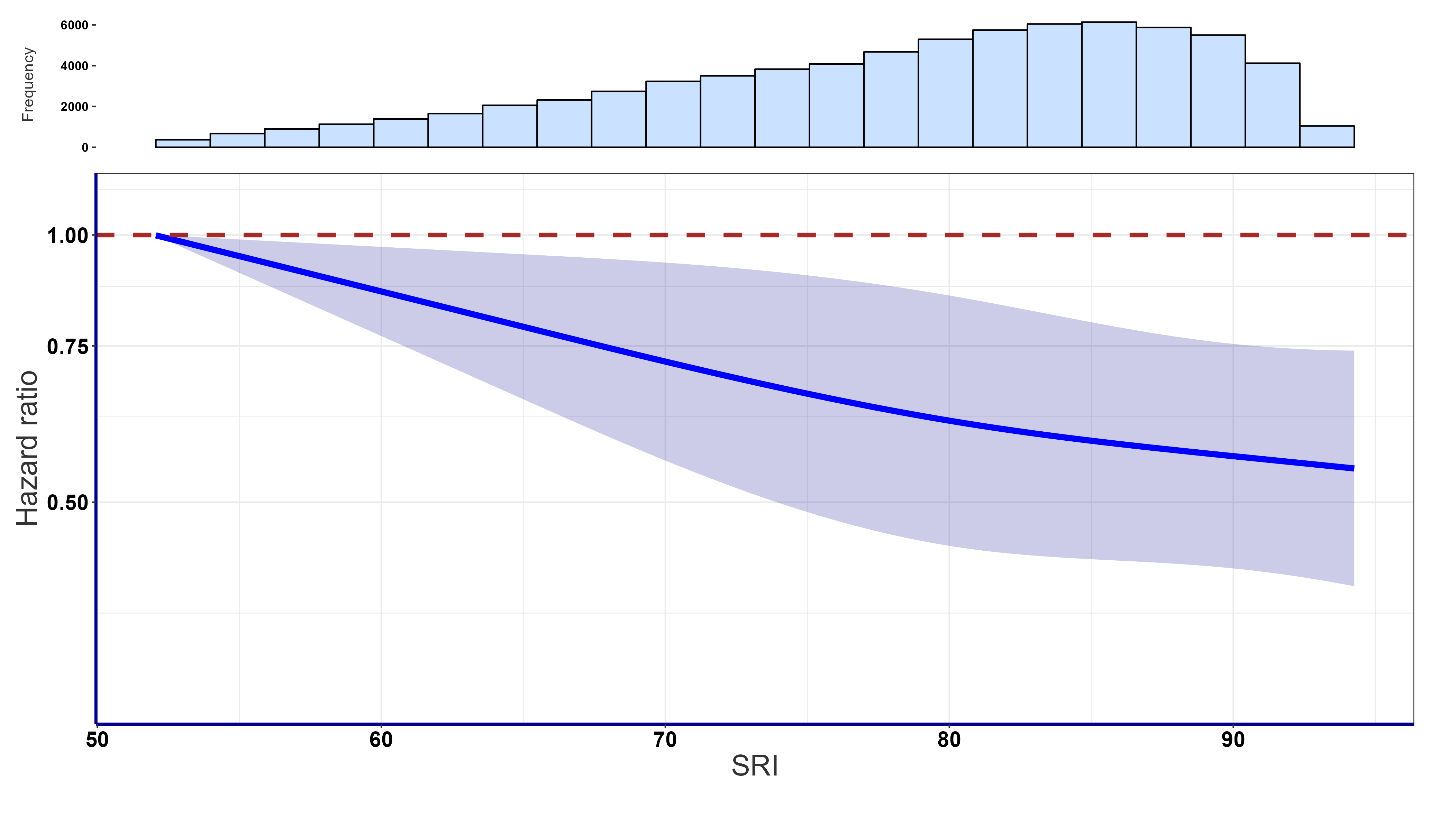

Dose-response curves showing incident dementia HR associated with increasing SRI. Reference point set to the lowest data point (SRI=51.5). Adjusted for age, sex, ethnic, BMI, fruit and vegetable consumption, smoking, alcohol consumption, coffee consumption, mental health issue, sleeplessness/insomnia, education, shiftwork, LPA, MVPA, sedentary behaviour. Data are shown for n=72347 with 703 events and with a mean follow-up of 7.9 (1.0) years.

**Supplementary Figure 4**: Association of sleep regularity with incident dementia stratified by sleep duration with participants employed in shift work excluded (n=72347, 703 events).

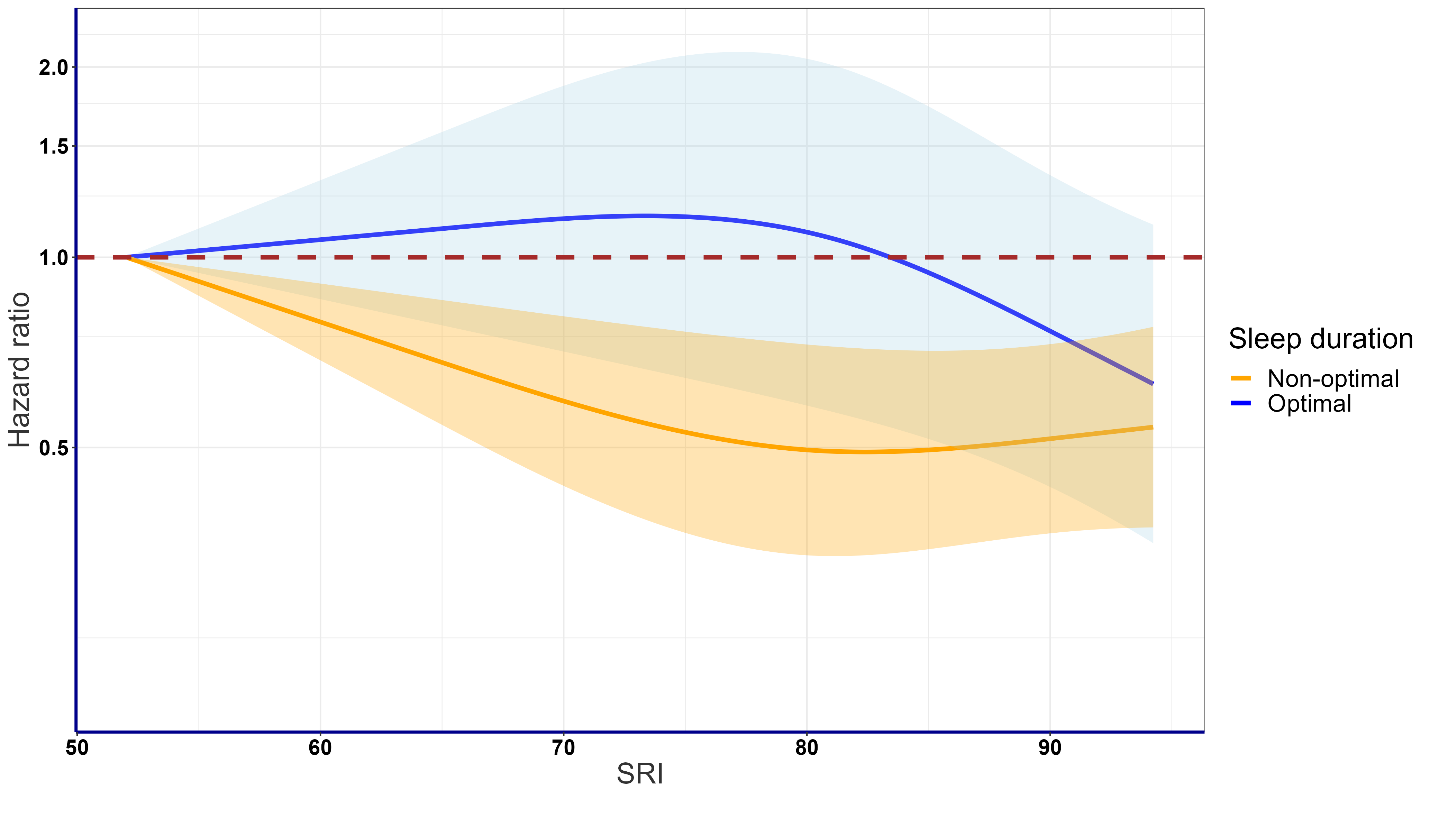

Dose-response curves showing incident dementia HR associated with increasing SRI stratified by two groups of sleep duration (ideal ≥ 7 and < 8h/day, and non-ideal <7h/day and ≥ 8h/day). Reference point set to lowest data point (SRI=51.0). Adjusted for age, sex, ethnic, BMI, fruit and vegetable consumption, smoking, alcohol consumption, coffee consumption, mental health issue, sleeplessness/insomnia, education, shiftwork, LPA, MVPA, sedentary behaviour. Data are shown for n=72347 with 703 events and with a mean follow-up of 7.9 (1.0) years.

**Supplementary Figure 5**: Association of sleep duration with incident dementia excluding participants with incident dementia in the first two years of follow-up (n=78197, 635 events).

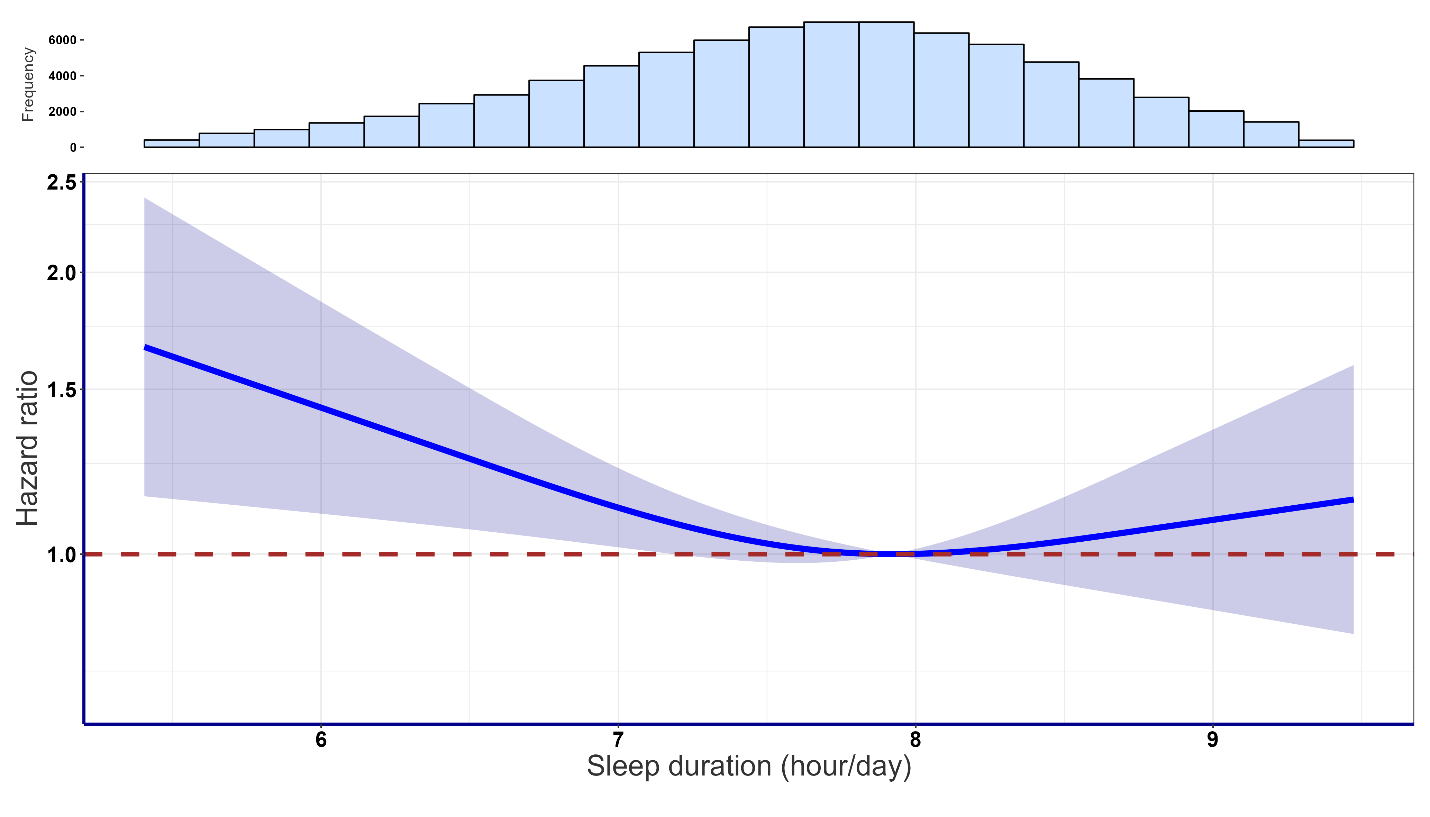

Dose-response curves showing incident dementia HR associated with increasing daily sleep duration. Reference point set to the optimal data point (7.9 hrs of sleep/day). Adjusted for age, sex, ethnic, BMI, fruit and vegetable consumption, smoking, alcohol consumption, coffee consumption, mental health issue, sleeplessness/insomnia, education, shiftwork, LPA, MVPA, Sedentary behaviour. Data are shown for n=78197 with 635 events and with a mean follow-up of 7.9 (1.0) years.

**Supplementary Figure 6**: Association of sleep regularity with incident dementia excluding participants with incident dementia in the first two years of follow-up (n= 78200, 667 events).

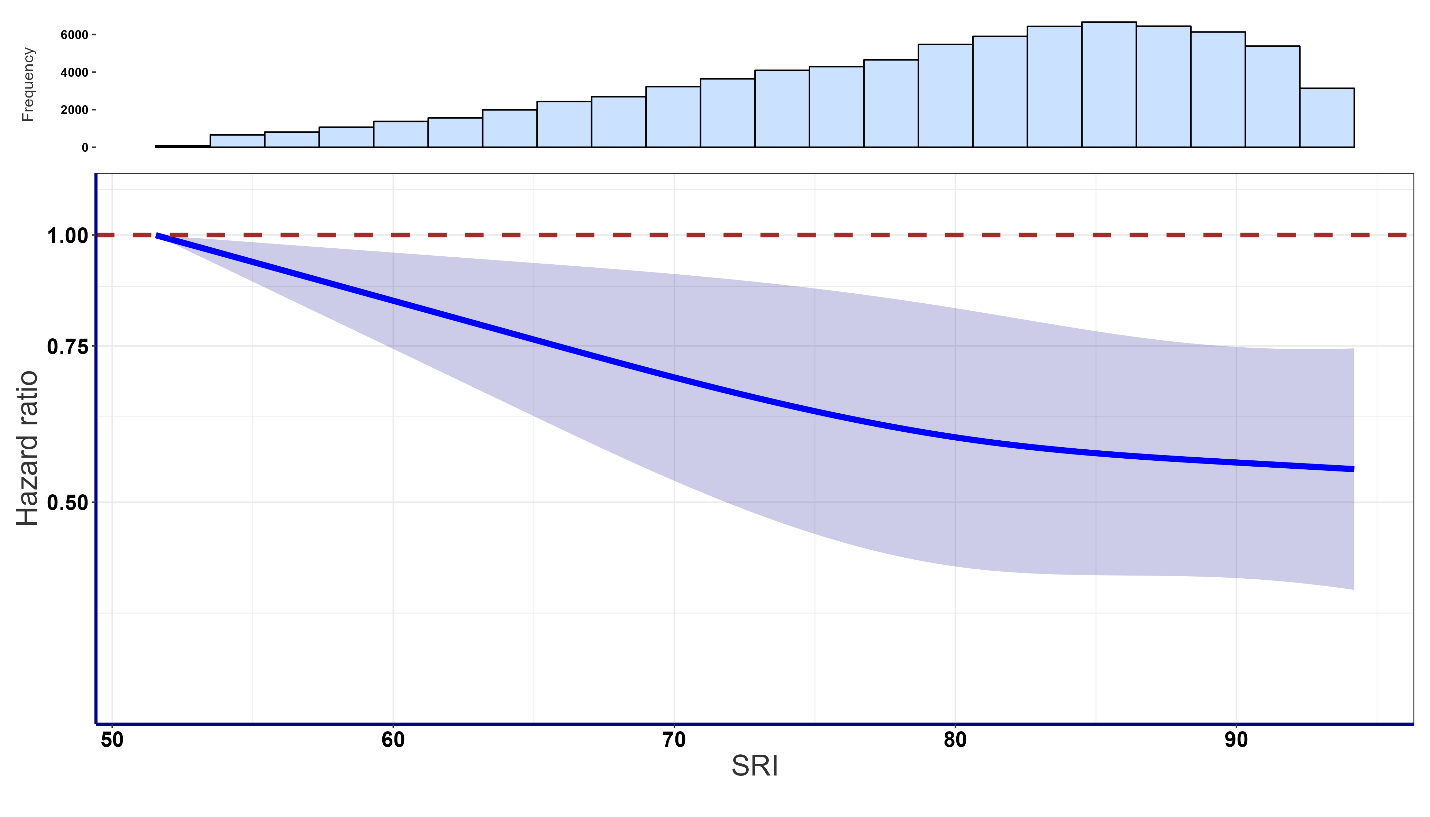

Dose-response curves showing incident dementia HR associated with increasing SRI. Reference point set to the lowest data point (SRI=51.5). Adjusted for age, sex, ethnic, BMI, fruit and vegetable consumption, smoking, alcohol consumption, coffee consumption, mental health issue, sleeplessness/insomnia, education, shiftwork, LPA, MVPA, sedentary behaviour. Data are shown for n=78200 with 667 events and with a mean follow-up of 7.9 (1.0) years.

**Supplementary Figure 7**: Association of sleep regularity with incident dementia stratified by sleep duration excluding participants with incident dementia in the first two years of follow-up (n= 78200, 667 events).

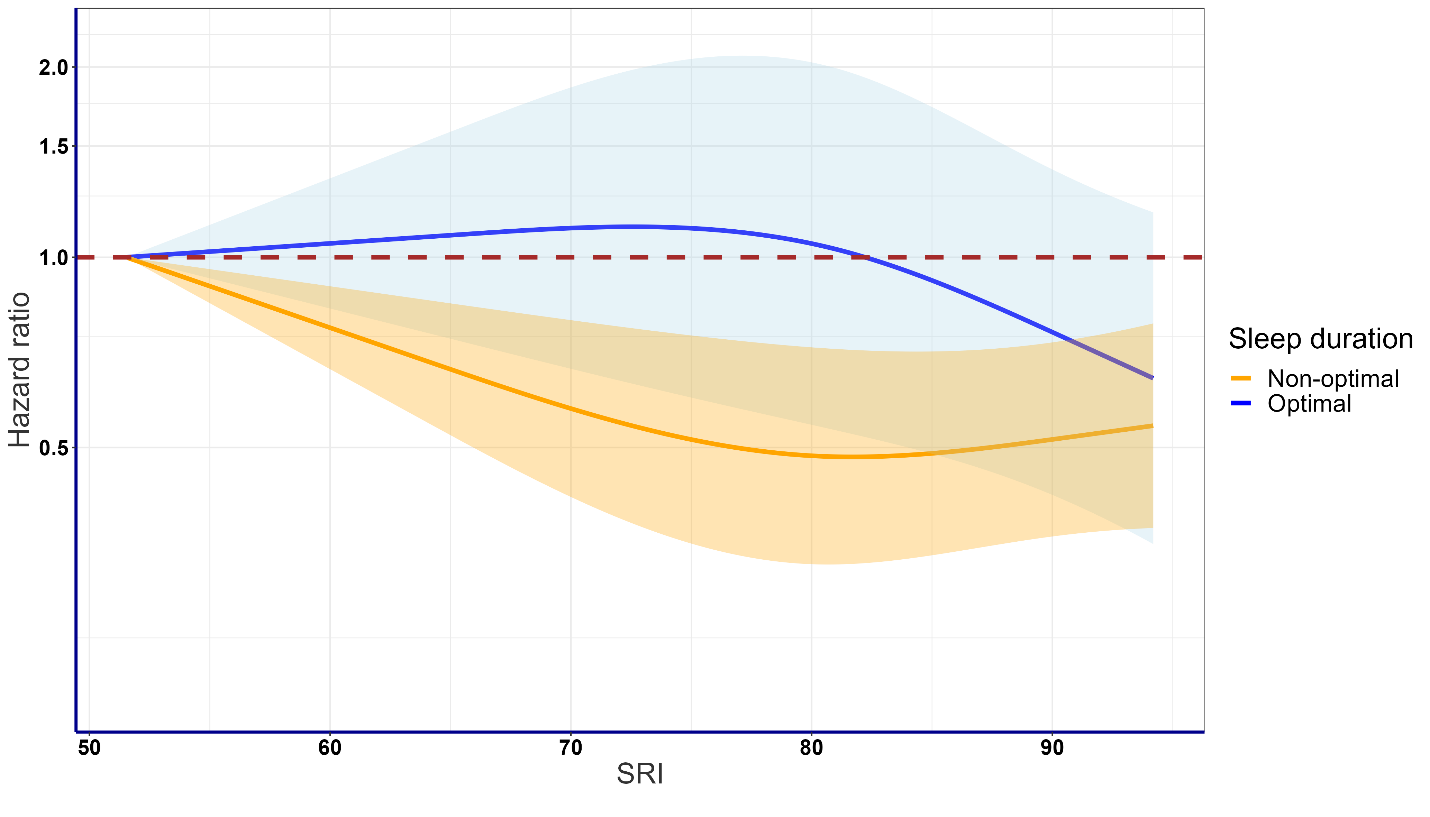

Dose-response curves showing incident dementia HR associated with increasing SRI stratified by two groups of sleep duration (ideal ≥ 7 and < 8h/day, and non-ideal <7h/day and ≥ 8h/day). Reference point set to lowest data point (SRI=51.5). Adjusted for age, sex, ethnic, BMI, fruit and vegetable consumption, smoking, alcohol consumption, coffee consumption, mental health issue, sleeplessness/insomnia, education, shiftwork, LPA, MVPA, sedentary behaviour. Data are shown for n=78200 with 667 events and with a mean follow-up of 7.9 (1.0) years.

**Supplementary Figure 8**: Association of sleep duration with incident dementia excluding participants with poor health (n=76311, 650 events).

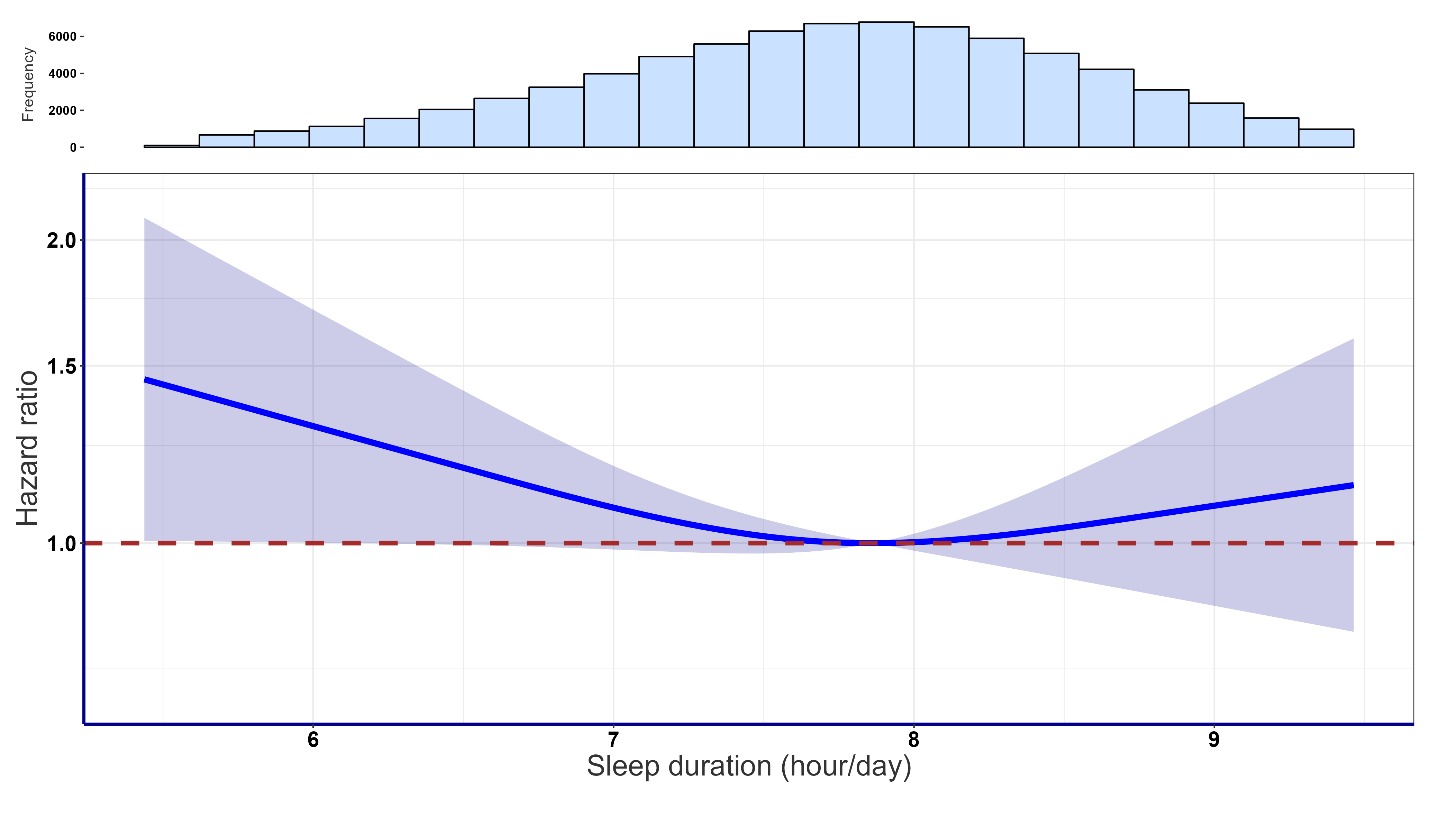

Dose-response curves showing incident dementia HR associated with increasing daily sleep duration. Reference point set to the optimal data point (7.9 hrs of sleep/day). Adjusted for age, sex, ethnic, BMI, fruit and vegetable consumption, smoking, alcohol consumption, coffee consumption, mental health issue, sleeplessness/insomnia, education, shiftwork, LPA, MVPA, Sedentary behaviour. Data are shown for n=76311 with 650 events and with a mean follow-up of 7.9 (1.0) years.

**Supplementary Figure 9**: Association of sleep regularity with incident dementia excluding participants with poor health (n= 76302, 681 events).

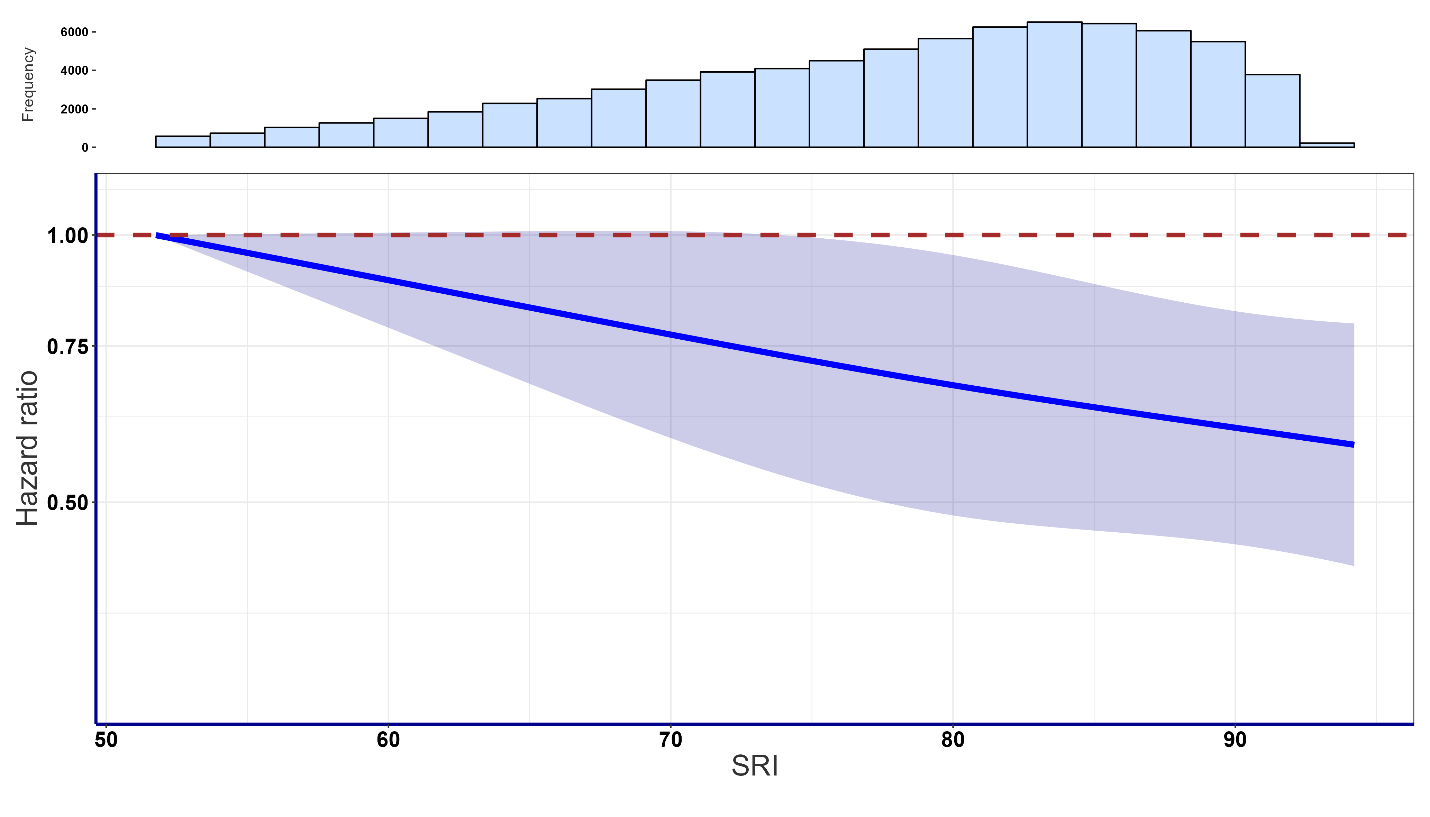

Dose-response curves showing incident dementia HR associated with increasing SRI. Reference point set to the lowest data point (SRI=51.7). Adjusted for age, sex, ethnic, BMI, fruit and vegetable consumption, smoking, alcohol consumption, coffee consumption, mental health issue, sleeplessness/insomnia, education, shiftwork, LPA, MVPA, sedentary behaviour. Data are shown for n=76302 with 681 events and with a mean follow-up of 7.9 (1.0) years.

**Supplementary Figure 10**: Association of sleep regularity with incident dementia stratified by sleep duration excluding participants with poor health (n= 76302, 681 events).

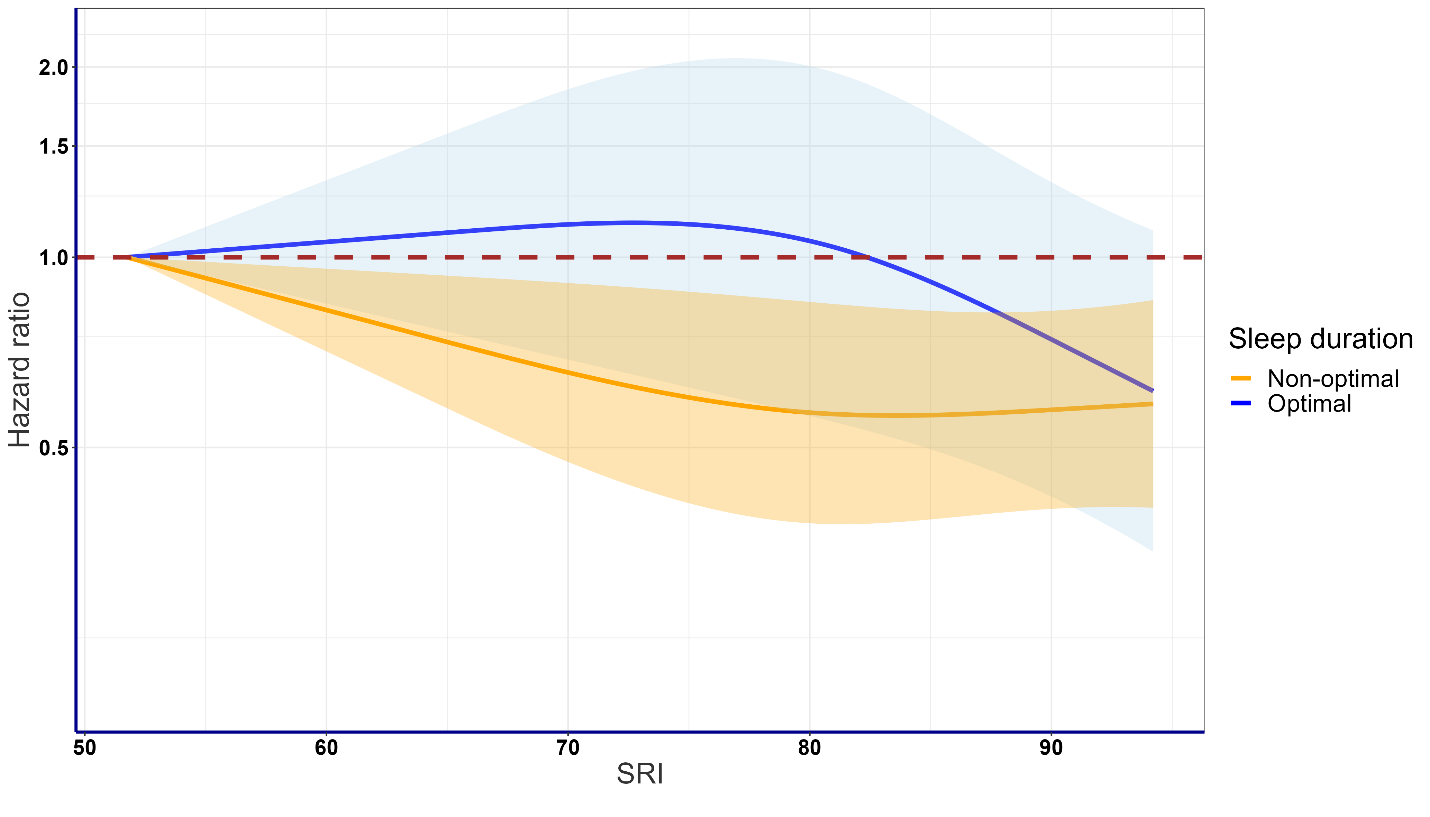

Dose-response curves showing incident dementia HR associated with increasing SRI stratified by two groups of sleep duration (ideal ≥ 7 and < 8h/day, and non-ideal <7h/day and ≥ 8h/day). Reference point set to lowest data point (SRI=51.7). Adjusted for age, sex, ethnic, BMI, fruit and vegetable consumption, smoking, alcohol consumption, coffee consumption, mental health issue, sleeplessness/insomnia, education, shiftwork, LPA, MVPA, sedentary behaviour. Data are shown for n=76302 with 681 events and with a mean follow-up of 7.9 (1.0) years.

**Supplementary Table 3:** Interaction between sleep duration and regularity

| **Interaction** | **P-value** |
| --- | --- |
| Duration*Regularity (minimal adjusted) | 0.6790 |
| Duration*Regularity (fully adjusted) | 0.8624 |

Interaction analysis of sleep duration and regularity with minimal adjustment for only age and sex, and with fully adjustment for all covariates. P-values represent the significance of the interaction terms between sleep duration and regularity.

**Supplementary Figure 11**: Association of categorised sleep duration with incident dementia n= 78256, 694 events).

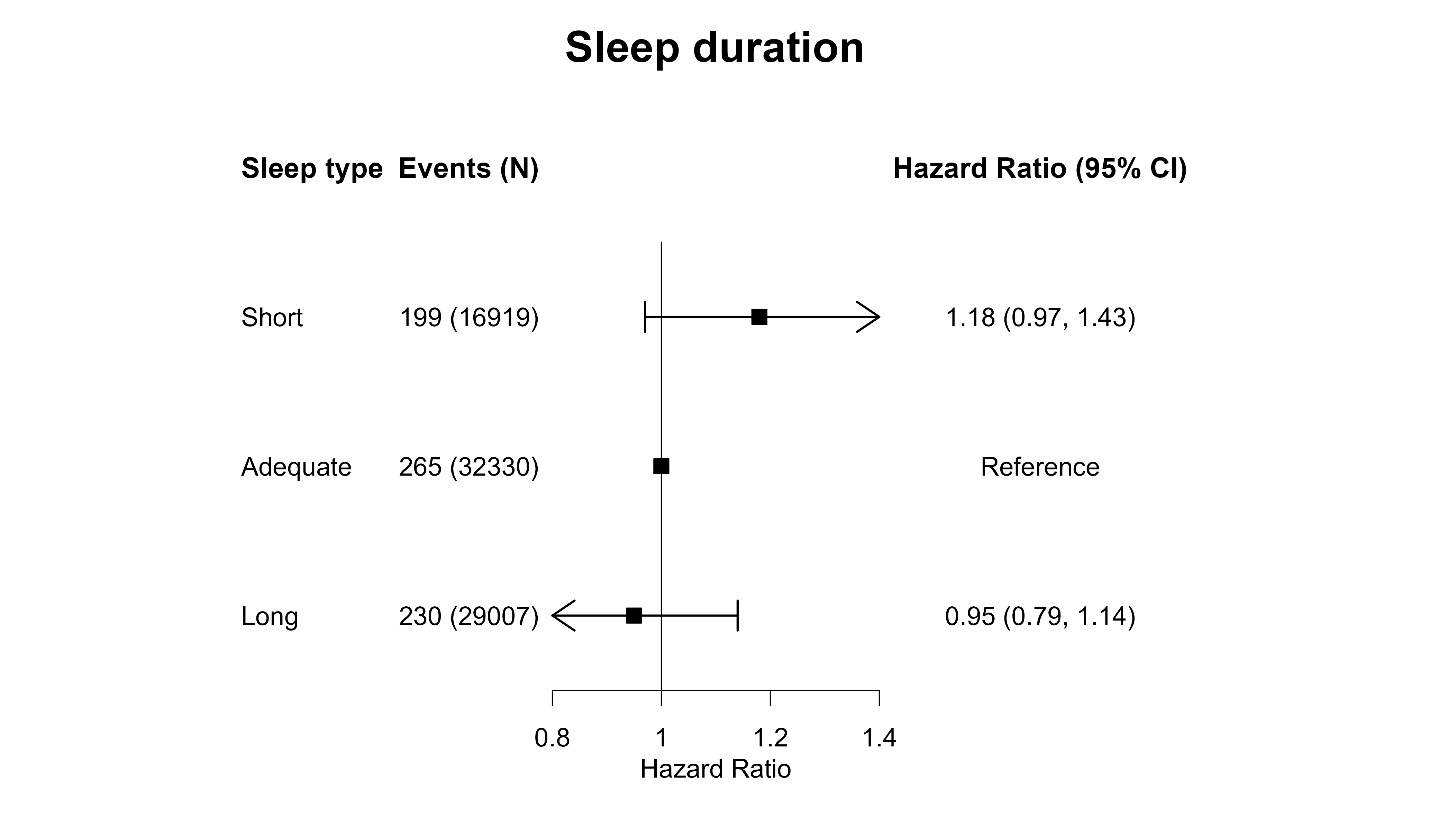

Forest plot showing incident dementia HR associated with daily sleep duration (short <7h/day, adequate ≥ 7 and < 8h/day, and long ≥ 8h/day). Reference group set to short sleep duration. Adjusted for age, sex, ethnic, BMI, fruit and vegetable consumption, smoking, alcohol consumption, coffee consumption, mental health issue, sleeplessness/insomnia, education, shiftwork, LPA, MVPA, sedentary behaviour. Data are shown for n=78256 with 694 events and with a mean follow-up of 7.9 (1.0) years. Error bars represent 95% CI.

**Supplementary Figure 12**: Association of categorised sleep regularity with incident dementia n= 78262, 729 events).

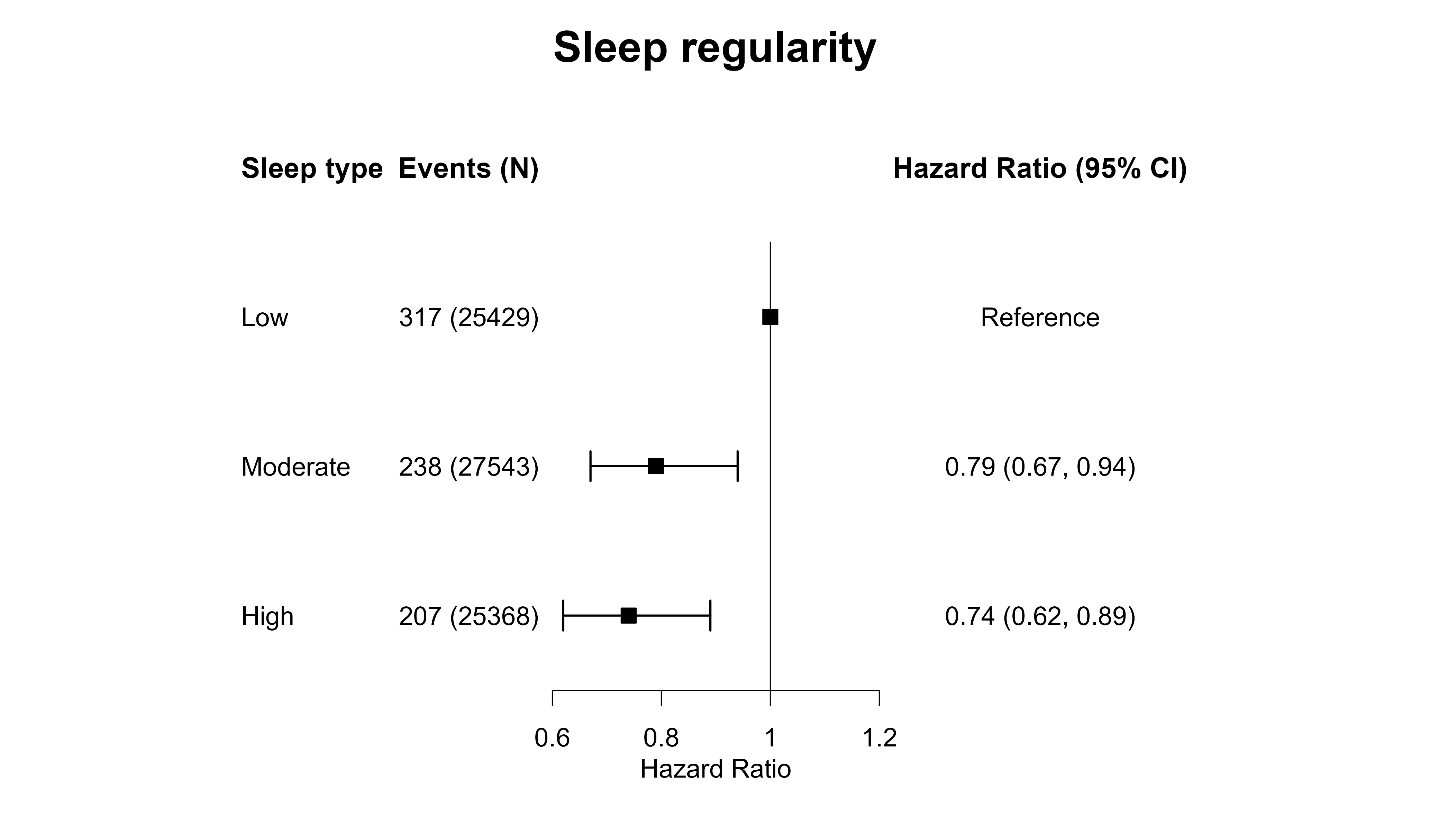

Forest plot showing incident dementia HR associated with daily sleep regularity (categorised based on tertiles of the SRI distribution, low <74.95, moderate ≥ 74.95 and < 85.05, and high ≥ 85.05). Reference group set to low sleep regularity. Adjusted for age, sex, ethnic, BMI, fruit and vegetable consumption, smoking, alcohol consumption, coffee consumption, mental health issue, sleeplessness/insomnia, education, shiftwork, LPA, MVPA, sedentary behaviour. Data are shown for n=78262 with 729 events and with a mean follow-up of 7.9 (1.0) years. Error bars represent 95% CI.
